# Modelling the impact of COVID-19-related programme interruptions on visceral leishmaniasis in India

**DOI:** 10.1101/2020.10.26.20219758

**Authors:** Epke A Le Rutte, Luc E Coffeng, Johanna Muñoz, Sake J de Vlas

## Abstract

**Background:** In March 2020, India declared a nationwide lockdown to control the spread of COVID-19. As a result, control efforts against visceral leishmaniasis (VL) were interrupted.

**Methods:** Using an established age-structured deterministic VL transmission model, we predicted the impact of a 6 to 24-month programme interruption on the timeline towards achieving the VL elimination target, as well as on the increase of VL cases. We also explored the potential impact of a mitigation strategy after the interruption.

**Results:** Delays towards the elimination target are estimated to range between 0 to 9 years. Highly endemic settings where control efforts have been ongoing for 5-8 years are most affected by an interruption, for which we identified a mitigation strategy to be most relevant. However, more importantly, all settings can expect an increase in the number of VL cases. This increase is substantial even for settings with a limited expected delay in achieving the elimination target.

**Conclusion:** Besides implementing mitigation strategies, it is of great importance to try and keep the duration of the interruption as short as possible, to prevent new individuals from becoming infected with VL, and continue the efforts towards VL elimination as a public health problem in India.

## Introduction

On the 25^th^ of March 2020, India declared a nationwide lockdown to control the spread of the COVID-19 pandemic.^1^ As a result, control programmes and preventive measures against many diseases were altered or interrupted, including national visceral leishmaniasis (VL) control efforts. VL is a protozoal infection transmitted by sand flies, which causes fever and eventually death when left untreated. It is controlled by indoor residual spraying of insecticide (IRS), as well as active case detection (ACD). According to WHO guidelines in response to the pandemic, the VL control programme had to suspend all activities, inevitably impacting the progress towards achieving the target of VL elimination on the Indian subcontinent (ISC).^2^

In the past decade, the incidence of VL has decreased drastically on the ISC, due to the implementation of successful control. The VL control strategy starts with a 5-year ‘attack phase’, which consists of intense IRS and ACD, followed by an ongoing ‘consolidation phase’ with less intense IRS but increased ACD. Elimination of VL as a public health problem is considered to be achieved when the VL incidence remains below 1 VL case per 10,000 people per year at sub-district level for 3 consecutive years.^3^ It is important to estimate the impact of halting control efforts in terms of the timeline towards achieving the VL elimination target and the extent at which the hard-won gains from previous years are lost. Furthermore, decision makers would like to know whether and how these effects could be mitigated with existing tools.

In this study, we quantify the potential impact of the interruption of the VL control programme in India, using an established deterministic age-structured VL transmission model. More specifically, we estimate both the delay in reaching the VL elimination target as well as the increase of true VL incidence (i.e. all cases, not only those detected). Further, we analyse the impact of a mitigation strategy, implemented as an extension or a re-introduction of the attack phase, of equal length as the duration of the interruption, to counter the losses caused by the interruption of the VL control programme.

## Methods

### Model structure and quantification

We employed the established age-structured deterministic VL transmission model described by Le Rutte *et al*.,^4–7^ which captures the transmission of VL between humans and sand flies on the Indian subcontinent. The model considers that most infected individuals are asymptomatic and recover without ever having symptoms; a small fraction (∼1.5%) of individuals become symptomatic and will require treatment or will die otherwise (only a small fraction (∼3%) of symptomatic cases recover spontaneously). Transmission is driven by exposure to sand flies, which can pick up infection from symptomatic cases and individuals with post-kala-azar dermal leishmaniasis (PKDL), a mostly self-limiting but long-lasting skin condition that occurs in a fraction (2.5%) of individuals treated for VL. In addition, we consider the possibility that asymptomatically infected individuals do (model E1) or do not (model E0) contribute to transmission. The model incorporates IRS coverage through a user-defined proportional reduction in the sand fly population density, and ACD through a decrease in the average detection delay of symptomatic cases (baseline 60 days). Model parameters were previously calibrated based on age-structured data from approximately 21,000 individuals included in the KalaNet bednet trial in India and Nepal.^5,9^ The impact of IRS was estimated using a geographical cross-validation on case incidence in Bihar (∼6,000 VL cases in 8 districts over a period of 18 months). ^5,9^ A schematic representation of the model structure is presented in Supplementary Information Figure S1. The model was coded in R (version 4.0.2), using the *pomp* package (version 3.1.1.7); the model code is publicly accessible at [GITLAB LINK]. We provided the Policy-Relevant Items for Reporting Models in Epidemiology of Neglected Tropical Diseases (PRIME-NTD) Summary in Table S1 which was established to set a standard and increase consistency among publications using modeling to inform policy.^10^

### Scenarios

We considered populations with different levels of pre-control VL incidence (both detected and undetected cases) ranging from 2 to 12 VL cases per 10,000 capita per year. During the pre-control phase we assume only passive case detection to be in place, leading to an average duration between the start of symptoms and the start of treatment of 60 days. The simulated control programme starts with a 5-year attack phase in which we assume IRS to reduce the number of sand flies in the model by 67% and ACD which reduces the time to treatment to an average of 45 days. Subsequently, during the consolidation phase, IRS coverage is reduced to 45%, but ACD efforts are further increased leading to an average duration to treatment of 30 days. We refer to this scenario as the counterfactual scenario (i.e. no interruption due to COVID-19 occurs).

Next, we defined a range of scenarios in which the aforementioned control strategy is interrupted by COVID-19 at various time points during the control program (at 0 to 15 years after initiation of control) and for various durations of interruption (6, 12, 18, and 24 months), leading to a total of 176 simulations per model, per duration of interruption (11 pre-control endemicity levels x 16 time points of interruption during the control programme). During an interruption, we assume that no IRS and ACD take place and that only passive case detection remains, similar to the pre-control scenario (i.e. an average treatment delay of 60 days). We further assume that the interruption does not change (1) the rate at which humans are exposed to sand flies or (2) the risk of developing PKDL and the duration of PKDL. After the interruption, we assume that a control program either follows the original planning of interventions (i.e. the “interruption scenario”), or that a mitigation strategy is implemented to counter the effects of the interruption (“mitigation scenario”). Here, we let mitigation efforts depend on when the interruption occurs. When the interruption takes place during the attack phase (i.e. the first five years of a program), the attack phase is extended by the same duration as that of the interruption. When the interruption occurs during the consolidation phase (i.e. after 5+ years of control), a temporary attack phase similar in length to the duration of the interruption is initiated after the interruption and before continuing with the consolidation phase. The additional implemented attack phase contains an intense IRS strategy (67% sand fly reduction) and an ACD leading to a duration of time to treatment of 45 days, as during the regular attack phase.

### Impact assessment

For each combination of pre-control endemicity, timing of interruption, and duration of interruption, we compared A) the counterfactual scenario to the interruption scenario, B) the interruption scenario to the mitigation scenario, and C) the counterfactual scenario to the mitigation scenario. We quantified the impact of the interruption of the VL control programme in terms of (1) the delay in reaching the elimination target (in years) and (2) the increase in the cumulative number of new VL cases (detected and undetected). The target is defined as 1 VL case (detected and undetected) per 10,000 people per year in the modelled population for 3 consecutive years. In previous modelling studies the time of elimination was defined at the first moment of reaching an incidence <1 VL case per 10,000 people per year, however in this study we decided to set this target after 3 years of being below the target incidence, the moment when a setting can be considered by WHO for achieving validation of elimination. In case the elimination target had already been met before the interruption but was lost due to the interruption, we defined the delay as the time between the start of the interruption and the moment when the target was met again after the interruption (i.e. based on the interruption scenario only). When comparing the scenarios for differences in cumulative incidence we compared the area under the curve from year 0 (start of the control programme) to year 30 (by which time the effect of the interruption has fully waned).

## Results

The predicted impact of an interruption of the VL elimination programme due to the COVID-19 pandemic on VL elimination in India varies widely between settings. It depends on the stage of the control program at which the interruption takes place, the duration of the interruption, and the pre-control endemicity of a setting (Figures 2 and 3 and Figures S5-S18). Figure 1 illustrates that the impact of a 1 (blue line) or 2-year interruption (red line) is largest when the interruption takes place during the attack phase, followed by the interruption taking place early in the consolidation phase, and that the impact is relatively minor when the interruption takes place late in the consolidation phase, for a setting with 9 VL cases/10,000/year pre-control.

**Figure 1.**
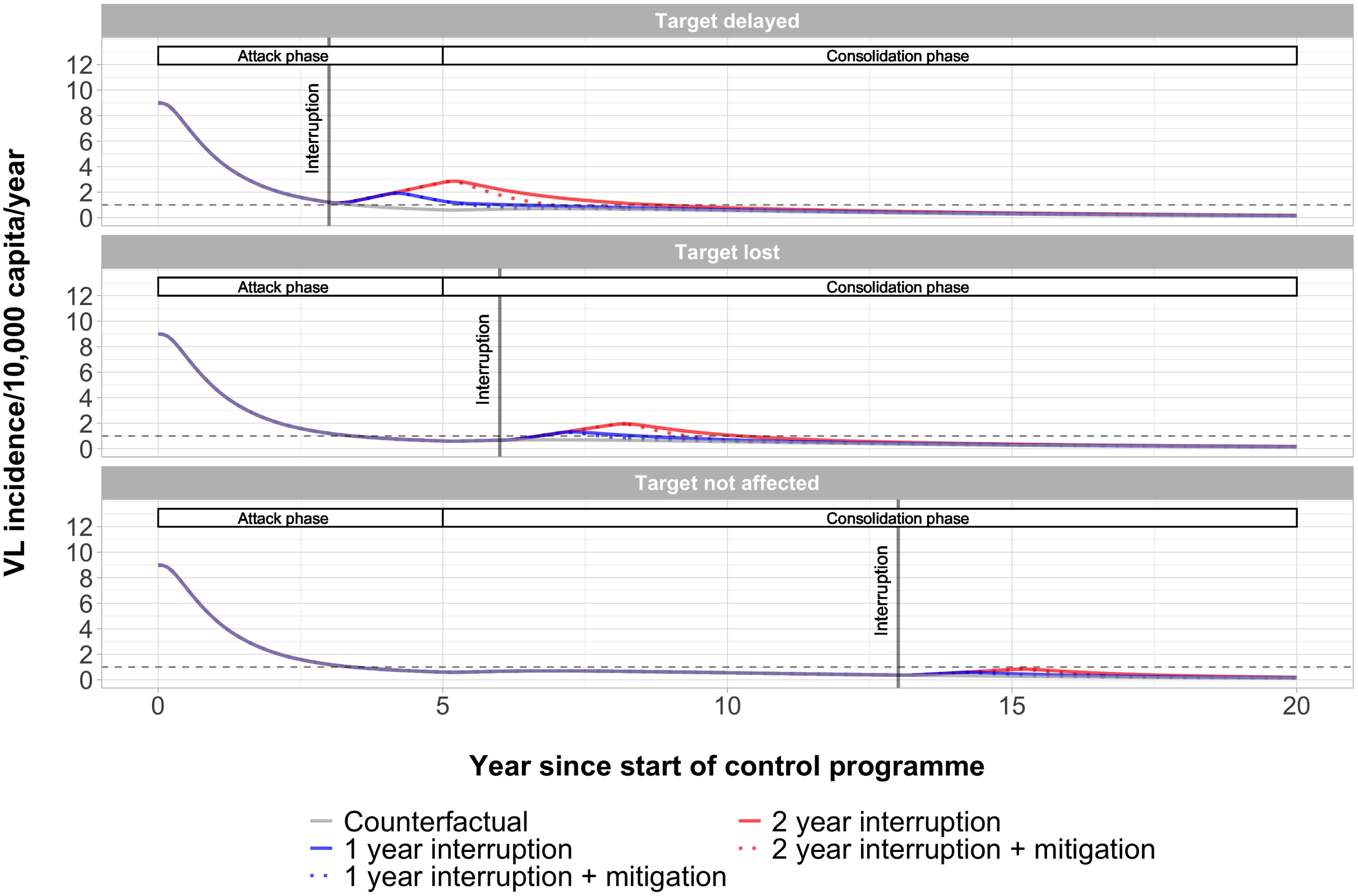
Predicted visceral leishmaniasis (VL) incidence over time by Model E1. Three interruption scenarios are presented for a setting with a pre-control endemicity of 9 VL cases/10,000/year. The white bars at the top of each panel stating ‘Attack phase’ and ‘Consolidation phase’ represent the course of the control strategy for the counterfactual scenario. Model E1 assumes that asymptomatic individuals contribute to transmission; see Figure S4 for similar predictions by Model E0.

**Figure 2.**
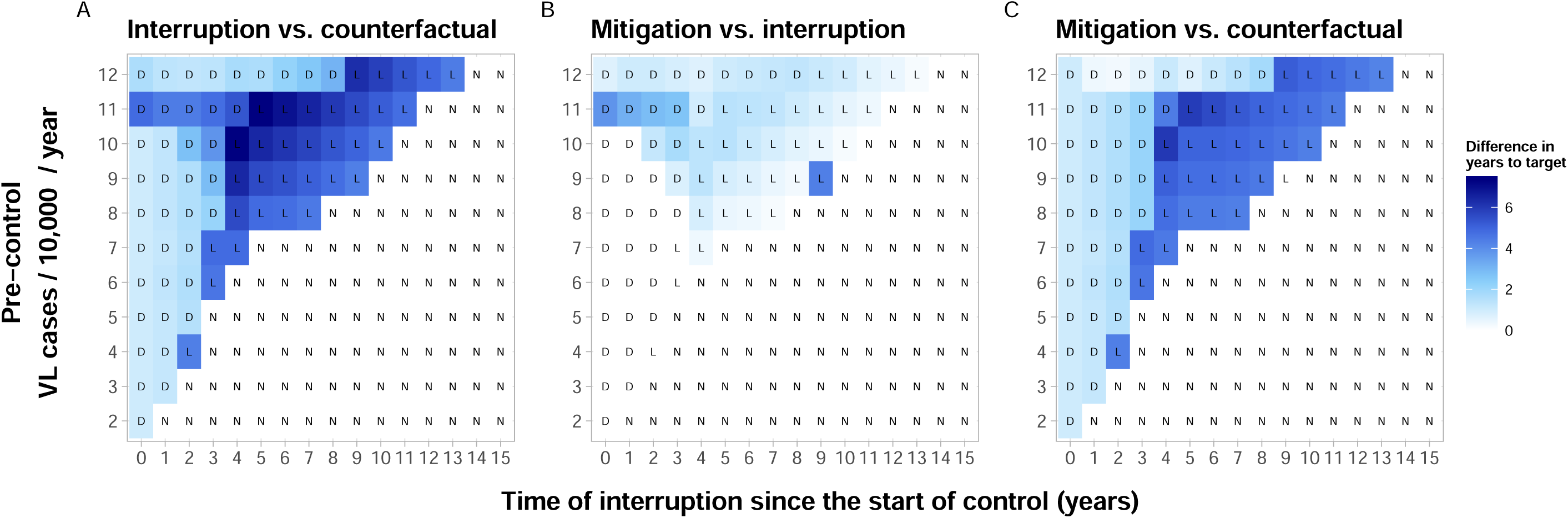
Heatmap of differences in time (years) to achieving the VL elimination target for three comparisons of scenarios using Model E1. Panel A: interruption vs. counterfactual (no interruption); Panel B: mitigation vs interruption; Panel C: mitigation vs counterfactual. Both interruption and mitigation are assumed to last one year. The letters D, L, and N correspond to the impact of the interruption on the target when comparing the counterfactual scenario to the interruption scenario; delayed (D), lost (L), or not affected (N), as illustrated in Figure 1. Results for Model E0 are shown in Figure S7.

**Figure 3.**
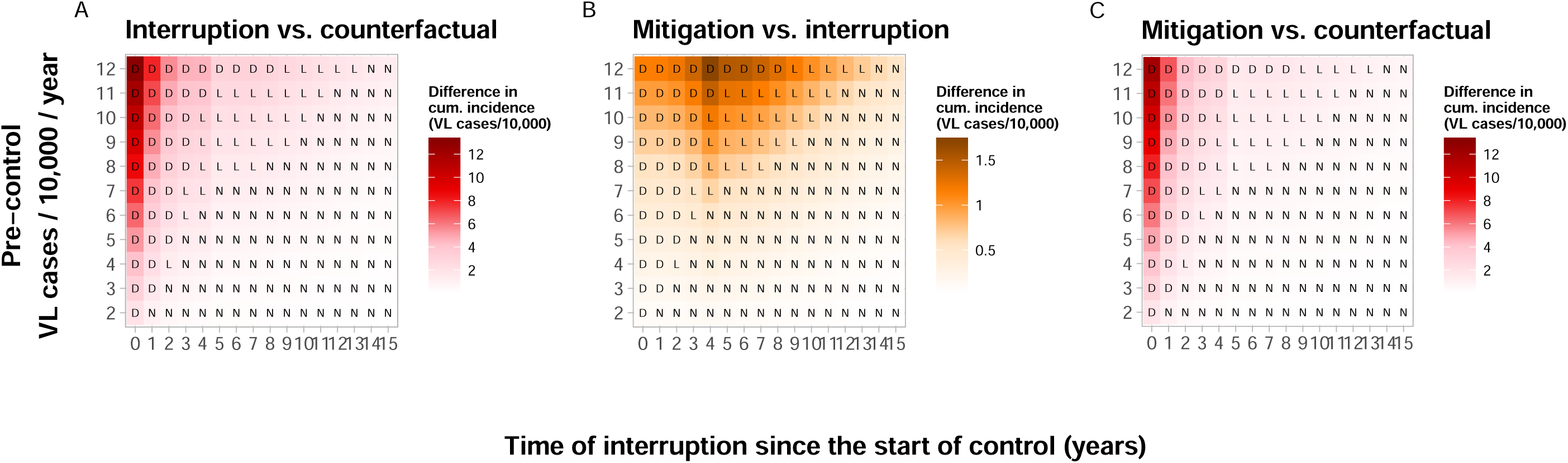
Heatmap for the difference in cumulative case incidence per 10,000 population for three comparisons of scenarios. Panel A: interruption vs. counterfactual; Panel B: mitigation vs interruption, and Panel C: mitigation vs counterfactual. We use a different colour in panel B to indicate the finer scale relative to that depicted in panels A and C. The letters D, L, and N correspond to the impact of a one-year interruption on the target when comparing the counterfactual scenario to the interruption scenario; delayed (D), lost (L), or not affected (N), as illustrated in Figure 1 (Model E0 in Figure S14).

With regard to the timing of the interruption, we can distinguish three types of outcomes in comparison to the counterfactual scenario (no interruption, grey line in Figure 1). First, there are settings which experience a delay in achieving the target due to the interruption (Figure 1, top panel). Secondly, there are settings in which the target was already achieved before the interruption took place, but due to the interruption the target is lost again (middle panel). Thirdly, there are settings in which the timing of achieving the target is not affected by the interruption (bottom panel). The time for achieving the target for all 11 pre-control counterfactual scenarios is presented in Figure S2 (Model E1) and Figure S3 (Model E0). In the counterfactual scenario for a setting with a pre-control incidence of 9/10,000/year, the target is achieved ∼6.5 years after the start of the programme. If the interruption takes place during the attack phase, three years after the start of the programme (top panel), a one-year interruption would lead to a delay of about 3 years and a two-year interruption to a delay of about 5.5 years. If the interruption takes place six years after the start of the programme, i.e. early in the consolidation phase, the achievement of the target would be lost (middle panel) for both the one and two-year interruption scenarios. It will take about 5 years or even 7 years, respectively, for the target to be achieved again, counting from the start of the interruption. If the interruption takes place thirteen years after the start of the programme, i.e. late in the consolidation phase, the timing of the achievement of the target would not be affected at all (bottom panel).

For the same setting with a pre-control endemicity of 9/10,000/year, the impact of a mitigation strategy was also found to vary between scenarios (dotted lines in Figure 1). If the target is delayed (top panel), a mitigation strategy could reduce this delay by nearly 1 year with a one-year interruption, or nearly 2 years with a two-year interruption. In a similar setting where the target is lost (middle panel), a mitigation strategy could reduce the delay by about half a year or 1 year, respectively. If the interruption occurs after 13 years of control (bottom panel), a mitigation strategy would obviously not provide benefit in terms of achieving the target. The outcomes of Model E0 were qualitatively similar and are presented in Figure S4.

In Figure 2 we provide an overview of the differences in delay to achieving the target for 176 scenarios (11 pre-control endemicity levels (y-axis) and 16 different stages of the control program at which interruption and mitigation takes place (x-axis)). In all underlaying simulations we assume the interruption and mitigation scenarios to last one year. The differences in years to achieving the target range from 0 to more than 7 years, where the delay is highest in a setting with a pre-control VL incidence of 10/10,000/year and an interruption at 4 years into the programme without mitigation (Figure 2A). In 40% of the settings the interruption leads to a delay or a loss of target with an average delay of 3.5 years. Generally, the settings most affected by an interruption are those with high pre-control endemicities, and when the interruption occurs between 4 and 9 years into the programme. Typically, for settings with lower pre-control incidences, the earlier into the programme the interruption takes place, the bigger the impact. Contrastingly, for settings with higher pre-control incidences, the later into the control programme, the bigger the impact. The mitigation strategy reduces the delays to the target by nearly 9 months on average (Figure 2B). In previously highly endemic settings (9— 12/10,000/year) this increases to an average of 13 months with a maximum of 7 years reduction. In moderately or low endemic settings, a mitigation strategy provides little additional impact as the target is estimated to be achieved around the same time with continued regular interventions. The outlier in Figure 2B (and Figures S6 and S7) is caused by the mitigation strategy leading to just not losing the elimination target whereas in the interruption scenario it does (Figure S19). Figure 2C shows that in no case the simulated mitigation strategy could lead to achieving the target sooner compared to the counterfactual scenario. Similar simulations with Model E0 (in which only VL and PKDL cases are considered infectious) follow an identical pattern (Figure S7). However, in 75% of the settings the interruption is estimated to lead to a delay and the impact occurs over the entire range of pre-control endemicities, but with a lower average delay of 2.5 years. For a setting with a pre-control incidence of 2/10,000/year, the delay is estimated to be as high as 6 years.

Would the interruption and potential mitigation last 6 months, fewer settings are estimated to be affected, 30% (Model E1) and 65% (Model E0) (Figures S5 and S6). The average delays are also expected to be lower, with an average of 2 years (E1) and 1.5 years (E0) delay and a maximum delay of ∼6.5 years (E1 and E0), about 6 months less compared to a 1-year interruption. However, would the interruption and potential mitigation last as long as 2 years, the impact would be larger, with about 55% (E1) and 80% (E0) of settings affected, an average delay of 5.5 years (E1) and 4 years (E0) in affected settings, and a maximum delay of 9 years (E1) and nearly 8 years (E0) (Figures S10 and S11). For all durations of interruptions, mitigation strategies are most impactful in the areas that are affected the most. The impact of an interruption and potential mitigation of 18 months is presented in Figure S8 (Model E1) and Figure S9 (Model E0), of which the impact lies in between the 1 and 2-year interruptions.

Besides the potential delays to achieving the elimination target we also predicted the impact of interruptions on cumulative VL incidence (detected and undetected cases) (Figure 3). All 176 simulated settings are affected by an increase in incidence due to the interruption (both Model E1 and E0), which ranged from 0.006—13/10,000 with an average of 1.8/10,000. The increase was highest in settings with the highest pre-control endemicity (i.e. 12/10,000/year among the simulated scenarios) and when the interruption taking place at the start of the programme (Figure 3 Panel A). This differs from the settings that are most affected based on their delay to the target (Figure 2 Panel A). A mitigation strategy could reduce the cumulative incidence with an average ∼1/10,000 in previously highly endemic settings (9—12/10,000/year). In moderately or low endemic settings, a mitigation strategy provides little additional impact (Figure 3 Panel B). A mitigation strategy can never fully compensate for previously increased incidence of VL, however it can bring incidence down faster compared to continuing with the regular control strategy, especially in high endemic settings that are 3 to 7 years into the programme. Similar simulations with Model E0 (in which only VL and PKDL cases are considered infectious) follow an identical pattern (Figure S14), however, with a higher average cumulative VL incidence of nearly 3/10,000 caused by the interruption.

After a 2-year interruption, the maximum cumulative incidence could be as high as 26/10,000 in settings with a pre-control scenario of 12/10,000/year with the interruption at the start of the control programme (Figure S17). This is twice as much compared to a 1-year interruption, while the delay to the target changes only from 7 years (1-year interruption) to 9 years (2-year interruption), which might seem less impactful. A 6-month interruption of the control programme (Figure S12), leads to half the cumulative incidence compared to a 1-year interruption, 50% less. However, when comparing a 1-year interruption to a 6-month interruption on the impact on the delay to the target, the difference is only 6 months (7 years and ∼6.5 years), 8% less. Therefore, a longer or shorter duration of the interruption might not have a major impact on the time of achieving the target, but highly impacts the number of additional VL cases.

## Discussion

The delay towards achieving the VL elimination target is predicted to vary widely between settings. A 1-year interruption of the VL control programme can cause 0 to 7 years of delay towards achieving the VL elimination target, compared to a situation without an interruption. In some settings that have already achieved the target previously to the COVID-19 pandemic, the target can be lost, as VL incidence increases to values above the target again. The length of this delay increases with pre-control endemicity, the duration of the interruption, and when the interruption happens within a few years of the switch from the attack to the consolidation phase of the programme. However, more importantly, an increase in VL incidence should be expected in all settings, and this increase is relatively speaking much higher than the expected delays. Mitigation strategies can in certain settings help to reduce the timeline towards the elimination target with as much as ∼4.5 years, and more importantly reduce the number of new VL cases; however, they can never make all losses caused by the interruption fully undone. Therefore, besides mitigation strategies, it is of great importance to try and keep the duration of the interruption as short as possible, to prevent new individuals from becoming infected with VL, to both positively impact the health of the population and reduce the delay to the VL elimination target.

As India’s sub-districts all have different pre-control endemicities and were in 2020 at different stages of the control programme, we simulated a range of 11 pre-control endemicities as well as 16 different years in which the interruption could take place (from the start of the control programme to 15 years into the programme). When interpreting the results, it is important to note that currently, about 80% of sub-districts in India are in the consolidation phase, mainly 5-10 years into the programme, of which the majority can be classified as previously moderately endemic settings (5-10/10,000/year). In our simulations we assume that both the IRS and ACD are implemented successfully during both the attack and consolidation phase. Further, it is important to note that our model predictions pertain to the total VL case incidence, including both detected and undetected cases. We previously showed that an increase in detection delay (be it due to scaling down control or a programme interruption) may result in a decline in reported VL cases, while transmission is already increasing again.^11^ Therefore, an evaluation of the actual impact of the COVID-19-related interruption of VL control should not only be based on reported VL cases, but also on the distribution of reported detection delays as this is an indicator of how many cases potentially remain undetected.

Besides the control target for VL on the Indian subcontinent, the global WHO VL 2030 target (85% of countries reaching <1% case fatality rate due to primary disease^12^) is also likely to be impacted by the global interruption of the programme due to COVID-19. With the predicted increases in VL incidence and the lack of active case detection and treatment during the interruption, an increase in VL-related mortality can also be expected. We did not present increased mortality rates as an outcome in this study, due to lack the data on mortality rates.

Although we aim to reflect the current situation as carefully possible, there are many real-life complexities that have changed during the lockdown period and have potentially influenced the transmission dynamics of VL, but that are not captured in the model. These include the massive population movement at the start of the lockdown, introducing many susceptible individuals from cities into endemic villages as well as introducing infected individuals into non-endemic villages. The lockdown then led to people staying more in and around their homes, which also may have led to changes in the contact rate between humans and sand flies and thus the transmission dynamics. Individual aspects such as increased malnutrition, and other co-morbidities that impact the course of infection of VL and associated infectiousness could also impact the transmission dynamics of VL adversely. Furthermore, we assumed that the care for and follow-up of detected VL cases (i.e. for detection and treatment of PKDL) would not be affected by the interruption. We previously showed that PKDL is an important driver of transmission after prolonged control.^7^ However, PKDL detection is likely to be impaired by COVID-19-related measures, as it relies on requires house visits by community healthcare workers. Because the duration of PKDL is already quite long (5 years in normal situations) compared to the duration of the interruption considered here (6-24 months), we expect that this assumption has little impact. Most likely, more PKDL cases will be detected and treated after the interruption such that the total average duration will remain approximately the same.

We have simulated a population at the scale of an Indian sub-district level using a deterministic age-structured model. Hence, no spatial heterogeneity or stochastic variation associated with finite populations was included. These factors are especially important in sub-districts with very low incidences as well as settings that are bordering sub-districts with different pre-control endemicities from their own. The latter however, might be less important when human movement is restricted. Furthermore, at a an even smaller geographical scale, such as village level, the potential effects of an interruption of control measures should be expected to be more variable, ranging from nothing to outbreaks as observed in Kosra.^13^ The latter was most likely influenced by the migration of people in and out of the area, a phenomenon that was also observed at the start of the Indian lockdown.

An important factor to consider is that the exact impact of both ACD as well as IRS on VL incidence remains debated. Even though their impact has been estimated by thoroughly fitting our models to data^5^, their direct impact still requires further investigation using more direct evidence. Given the uncertainty about the impact of COVID-19 on the control programmes, we decided to apply the pessimistic assumption of a complete withdrawal of all ACD and IRS activities. However, would for example still some IRS take place, or if ASHAs are still able to identify some individuals with fever and refer them to the community hospitals for VL diagnosis, the impact of COVID-19-related programme interruption would be smaller than predicted here.

Our results can at most be interpreted as a preview into the potential impact of the COVID-19 pandemic on VL in the ISC. Understanding the impact of the lockdown and other associated COVID-19 control measures on delays towards the VL elimination target and increased VL incidence will hopefully increase awareness for the additional health loss suffered by the areas that still experience VL. We wish to highlight that we should not lose focus of the VL elimination targets, that previous gains will to some extent be lost, but that introducing mitigation strategies and restarting the control programme as soon as it is again safe and possible, fortunately could reduce (but not counter) the losses. Resulting in the continuation of our efforts towards VL elimination as a public health problem on the ISC.

## Data Availability

No external datasets were used for this study, all data used to inform parameter values are open access and referenced in Supplementary Information Table 1.

## Authors’ contributions

EALR conceived the study; EALR and SJDV designed the study protocol; EALR and JM carried out the model simulations, EALR and LEC analysed and interpreted the outcomes. EALR drafted the manuscript; LEC, and SJDV critically revised the manuscript for intellectual content. All authors read and approved the final manuscript.

## Ethical approval

Not required

