## Supplementary Information for "Modelling the impact of COVID-19-related programme interruptions on visceral leishmaniasis in India"

#### 1. Table of Contents

**1. TABLE OF CONTENTS ... 1**
**2. MODEL STRUCTURE ... 2**
**3. MODEL PARAMETERS ... 2**
**4. PRIME-NTD SUMMARY TABLE ... 4**
**5. ADDITIONAL FIGURES ... 5**
5.1 PREDICTED VL INCIDENCE OVER TIME FOR COUNTERFACTUAL SCENARIOS ... 5
5.2 PREDICTED VL INCIDENCE OVER TIME FOR THREE OUTCOME SCENARIOS ... 6
5.3 DIFFERENCES IN TIME TO ACHIEVING VL ELIMINATION FOR 3 ALTERNATIVE DURATIONS OF INTERRUPTION ... 7
5.4 DIFFERENCES IN CUMULATIVE VL INCIDENCE FOR 3 ALTERNATIVE DURATIONS OF INTERRUPTION ... 9
**5. REFERENCES ... 12**

#### 2. Model structure

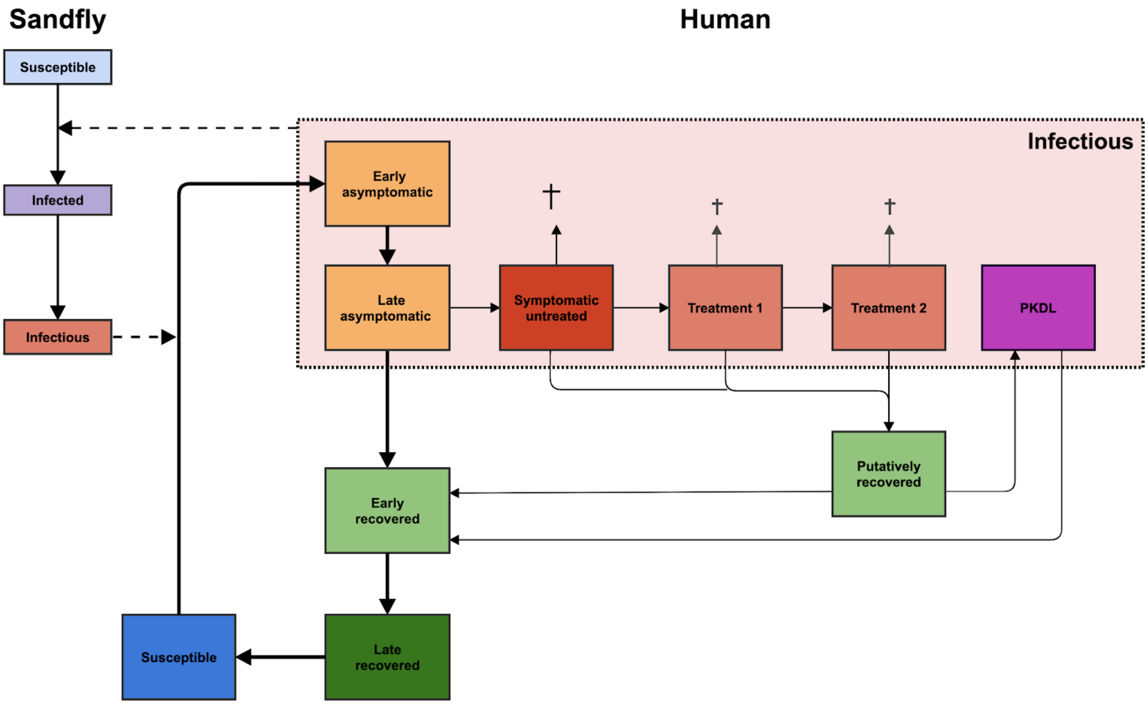

**Figure S1. Schematic presentation of the structure of model E1 and the related model E0.** For model E1, asymptomatic individuals (yellow compartments) are the main contributors to transmission. Model E0 has the same structure as model E1, but asymptomatic individuals do not contribute to transmission. Both models have different durations of infection stages from fitting to data, which are listed below in Chapter 3 Model parameters.

#### 3. Model parameters

**Table S1. Parameter values**

**Table 1. Overview of parameter values and assumptions**

| Parameters | Value[a] | Source |
| --- | --- | --- |
| Human birth rate (per 1000 capita) | 21 (Indian crude birth rate in 2011) | [1] |
| Human mortality rate | Age-dependent (Indian mortality rates in 2011) | [2] |
| Average duration of early asymptomatic stage (days) | 202 | Fitted to data in [3,4] |
| Average duration of late asymptomatic stage (days) | 69 | Fitted to data in [3,4] |
| Average duration of symptomatic untreated stage (days) | 60 (pre-control), 45 (attack-phase), 30 (consolidation phase) | [4–6] |
| Average duration of symptomatic treatment 1 (days) | 1 | [7] (CHECK) |
| Average duration of symptomatic treatment 2 (days) | 28 | [4,5,8] |
| Average duration of putatively recovered stage (months) | 21 | [9–11] |
| Average duration of PKDL (years) | 5 | Expert opinion and [10] |
| Average duration of early recovered stage (days) | 236 | Fitted in [3,4] |
| Average duration of late recovered stage (years) | 2 | Assumption based on [3] |
| Relative infectiveness of early asymptomatic individuals | 0.0114 (Model E1)<br>0 (Model E0) | Fitted (E1)<br>Pre-set (E0) |
| Relative infectiveness of late asymptomatic individuals | 0.0229(Model E1)<br>0 (Model E0) | Fitted (E1)<br>Pre-set (E0) |
| Relative infectiveness of symptomatic untreated cases | 1 | Reference value |
| Relative infectiveness of patients under treatment 1 and 2 | 0.5 | Expert opinion and [3] |
| Relative infectiveness of PKDL cases | 0.9 | [12,13] |
| Fraction of late asymptomatic individuals that become symptomatic untreated | 1.4% | Fitted in [3,4] |
| Fraction of untreated symptomatic cases that spontaneously, putatively recover | 3% | [14] |
| Excess mortality rate among untreated symptomatic cases (per day) | 1/150 | Assumption |

| | | |
|---|---|---|
| Excess mortality rate among treated symptomatic cases (per day) | 1/120 | Assumption [7,8] |
| Fraction of failed first-line treatments | 11% | Based on data presented in Supplementary File 2 of [4] |
| Fraction of putatively recovered cases that develop PKDL | 2.5 | [4,15,16] |
| Average life expectancy of the sandfly (days) | 14 | [17,18] |
| Average duration of incubation period in sandflies (days) | 5 | [19] |
| Sandfly biting rate (per day) | 0.25 | [20,21] |
| Transmission probability sandfly to human | 1.0[b] | Reference value |

[a] The parameter values listed here are the same for Models E0 and E1, unless stated otherwise.
[b] The probability that a susceptible person becomes infected when bitten by an infectious sandfly is assumed to be 1; potential overestimation is compensated by the estimated sandfly density per human.

#### 4. PRIME-NTD Summary Table

**Table S2. Policy-Relevant Items for Reporting Models in Epidemiology of Neglected Tropical Diseases (PRIME-NTD) Summary Table.** [22]

| Principle | What has been done to satisfy the principle? | Where in the manuscript is this described? |
|---|---|---|
| 1. Stakeholder engagement | Involved WHO HQ and PATH India | Introduction |
| 2. Complete model documentation | Described in detail in previous open access publications and on Github | Referred to previous papers in Methods, link to full open access of model code and documentation on Github in methods and here: [GITHUB LINK]. |
| 3. Complete description of data used | Described in detail in previous publications | Referred to particular datasets and previous papers in Methods[3,4] |
| 4. Communicating uncertainty | Described in detail in previous publications and also highlighted in this paper | Methods[4]/discussion |
| 5. Testable model outcomes | Not yet, in the future the model predictions can be compared to | Discussion |

| | |
|---|---|
| | KAMIS data. However, there is a discrepancy to be expected between real incidence and detected incidence. |

### 5. Additional figures

##### 5.1 Predicted VL incidence over time for counterfactual scenarios

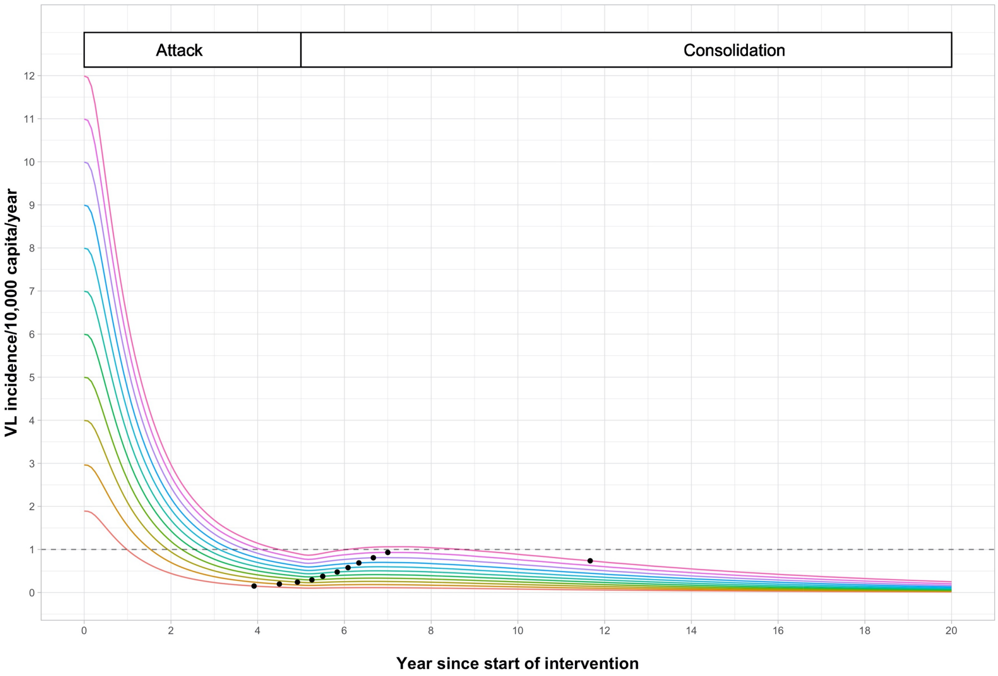

**Figure S2. Predicted visceral leishmaniasis incidence over time with expected times of achieving elimination (Model E1).** VL incidence is considered to be true incidence, so both detected and undetected cases. The coloured lines each represent a VL transmission setting with a different pre-control endemicity level. The white bars at the top stating 'Attack phase' and 'Consolidation phase' represent the course of the control strategy. The black dots represent the time of achieving elimination, which is defined as a VL incidence below 1 VL case per 10,000 people per year at sub-district level for 3 consecutive years.

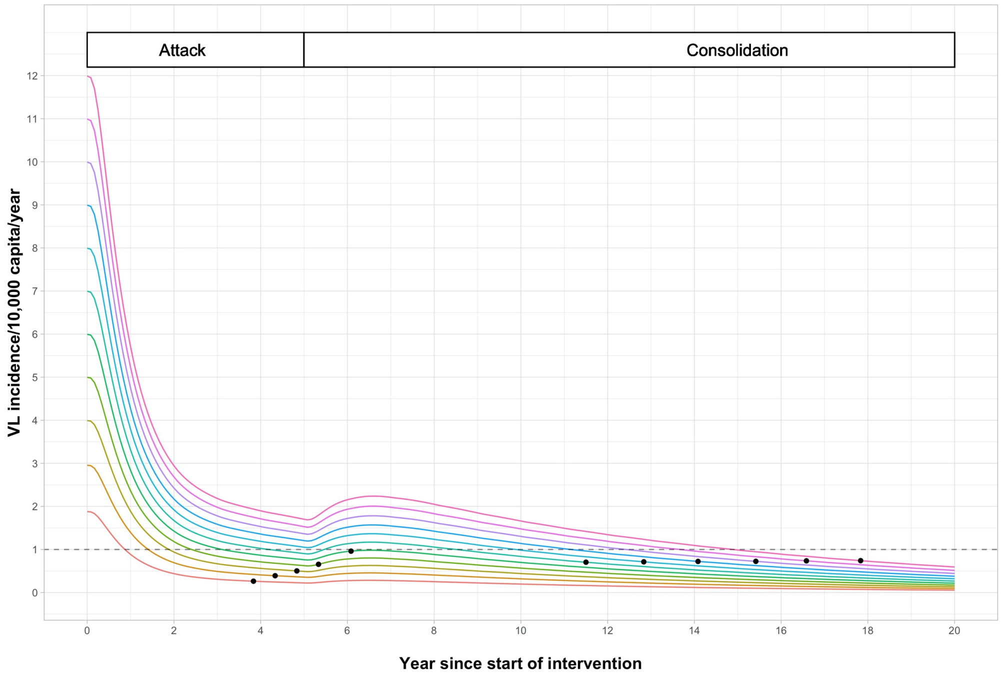

**Figure S3. Predicted visceral leishmaniasis incidence over time with expected times of achieving elimination (Model E0).** VL incidence is considered to be true incidence, so both detected and undetected cases. The coloured lines each represent a VL transmission setting with a different pre-control endemicity level. The white bars at the top stating 'Attack phase' and 'Consolidation phase' represent the course of the control strategy. The black dots represent the time of achieving elimination, which is defined as a VL incidence below 1 VL case per 10,000 people per year at sub-district level for 3 consecutive years.

#### 5.2 Predicted VL incidence over time for three outcome scenarios

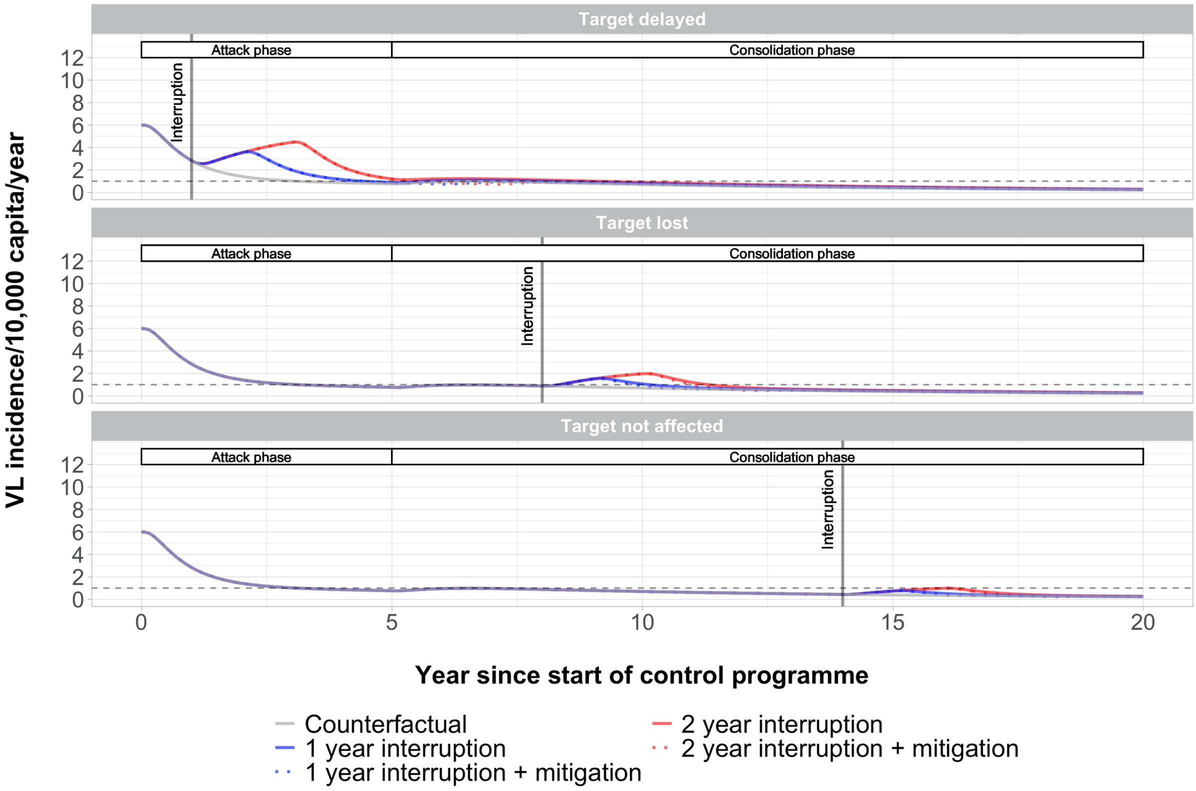

**Figure S4. Predicted visceral leishmaniasis incidence over time by Model E0.** Three interruption scenarios are presented for a setting with a pre-control endemicity of 6 VL cases/10,000/year. The white bars at the top stating 'Attack phase' and 'Consolidation phase' represent the course of the control strategy for the counterfactual scenario.

##### 5.3 Differences in time to achieving VL elimination for 3 alternative durations of interruption

The outliers that are present in Panel B of Figures SI-6, and SI-7 as well as in Figure 2 from the main text with are caused by the mitigation strategy leading to just not losing the elimination target whereas in the interruption scenario it does.

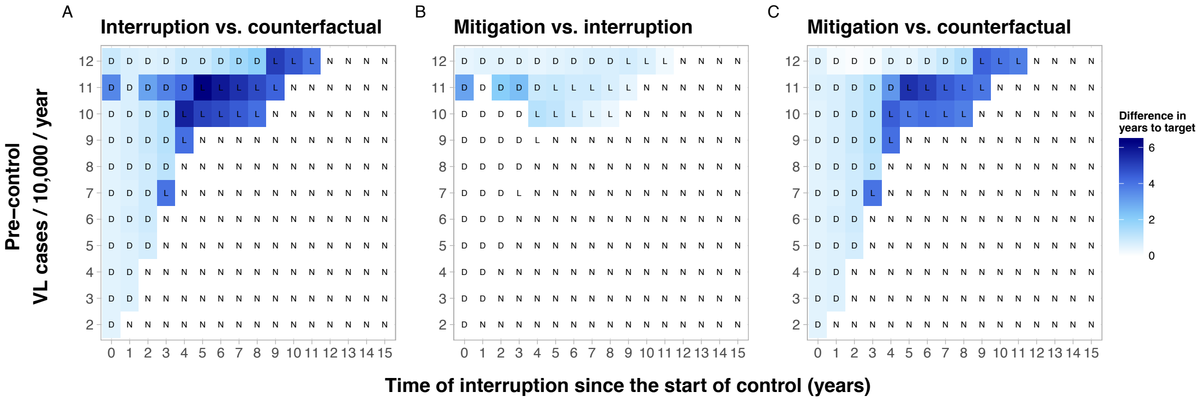

**Figure S5. Model E1: Heatmaps presenting the differences in time to achieving the VL elimination target for three combinations of scenarios (in years) with a 6-month interruption.** Panel A: interruption vs. counterfactual; Panel B: mitigation vs interruption, and Panel C: mitigation vs counterfactual. The letters D, L, and N correspond to the impact on the target when comparing the counterfactual scenario to the interruption scenario; delayed, lost, or not affected.

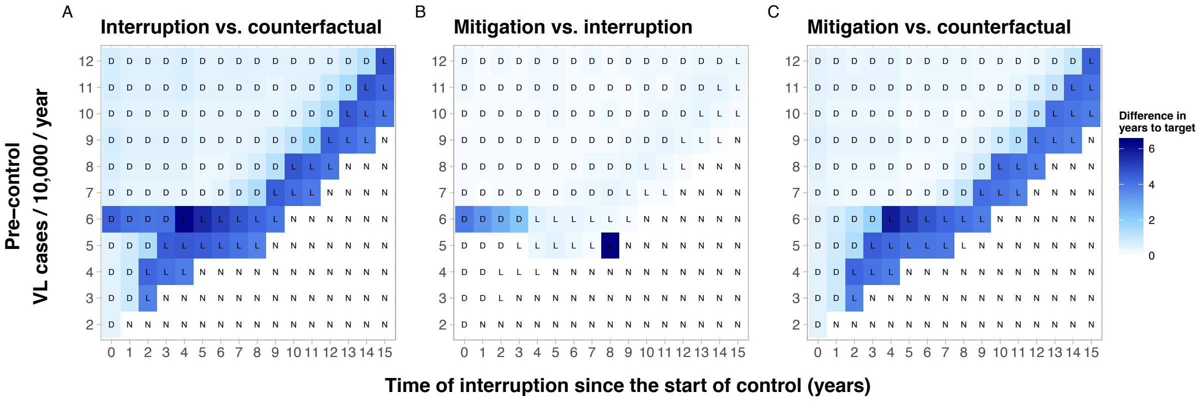

**Figure S6. Model E0: Heatmaps presenting the differences in time to achieving the VL elimination target for three combinations of scenarios (in years) with a 6-month interruption.** Panel A: interruption vs. counterfactual; Panel B: mitigation vs interruption, and Panel C: mitigation vs counterfactual. The letters D, L, and N correspond to the impact on the target when comparing the counterfactual scenario to the interruption scenario; delayed, lost, or not affected.

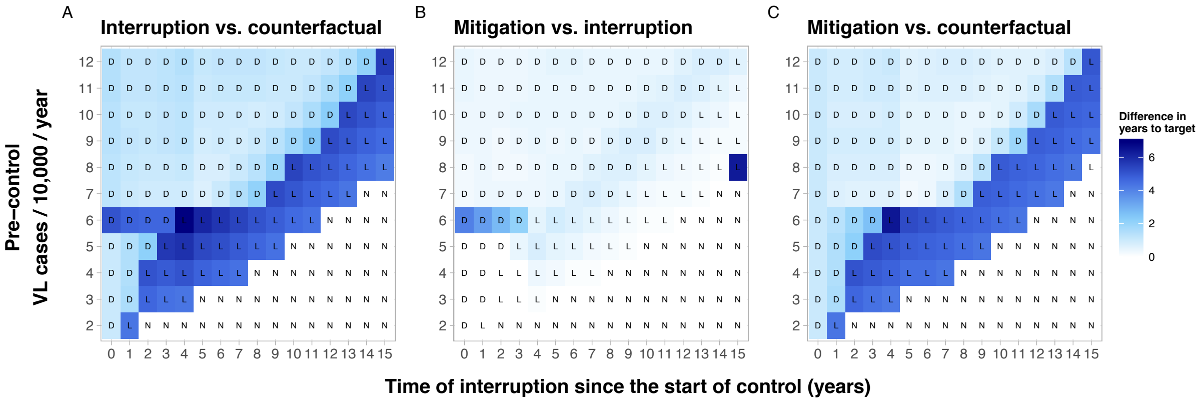

**Figure S7. Model E0: Heatmaps presenting the differences in time to achieving the VL elimination target for three combinations of scenarios (in years) with a 12-month interruption.** Panel A: interruption vs. counterfactual; Panel B: mitigation vs interruption, and Panel C: mitigation vs counterfactual. The letters D, L, and N correspond to the impact on the target when comparing the counterfactual scenario to the interruption scenario; delayed, lost, or not affected.

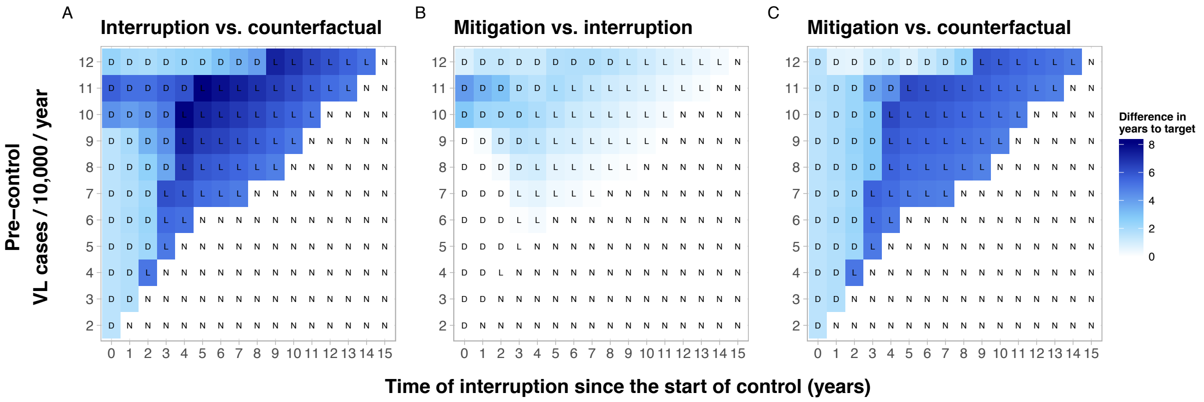

**Figure S8. Model E1: Heatmaps presenting the differences in time to achieving the VL elimination target for three combinations of scenarios (in years) with an 18-month interruption.** Panel A: interruption vs. counterfactual; Panel B: mitigation vs interruption, and Panel C: mitigation vs counterfactual. The letters D, L, and N correspond to the impact on the target when comparing the counterfactual scenario to the interruption scenario; delayed, lost, or not affected.

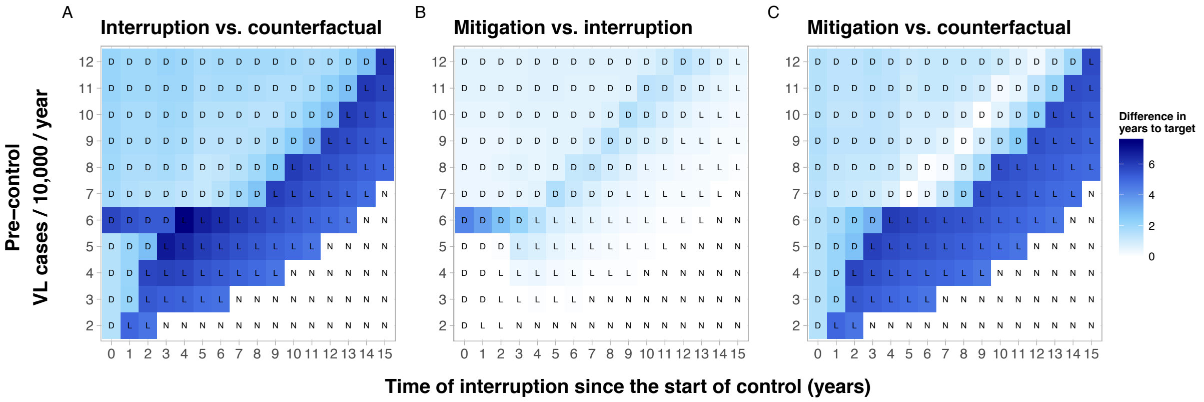

**Figure S9. Model E0: Heatmaps presenting the differences in time to achieving the VL elimination target for three combinations of scenarios (in years) with an 18-month interruption.** Panel A: interruption vs. counterfactual; Panel B: mitigation vs interruption, and Panel C: mitigation vs counterfactual. The letters D, L, and N correspond to the impact on the target when comparing the counterfactual scenario to the interruption scenario; delayed, lost, or not affected.

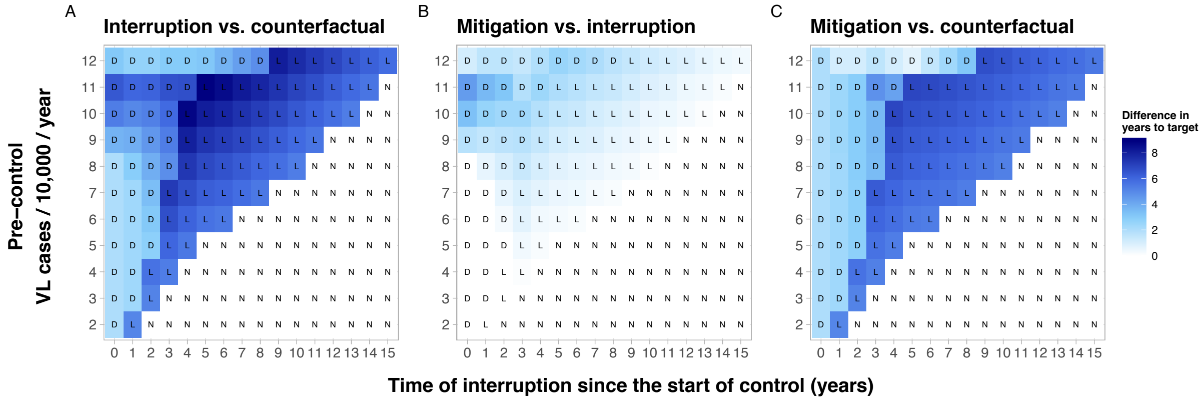

**Figure S10. Model E1: Heatmaps presenting the differences in time to achieving the VL elimination target for three combinations of scenarios (in years) with a 24-month interruption.** Panel A: interruption vs. counterfactual; Panel B: mitigation vs interruption, and Panel C: mitigation vs counterfactual. The letters D, L, and N correspond to the impact on the target when comparing the counterfactual scenario to the interruption scenario; delayed, lost, or not affected.

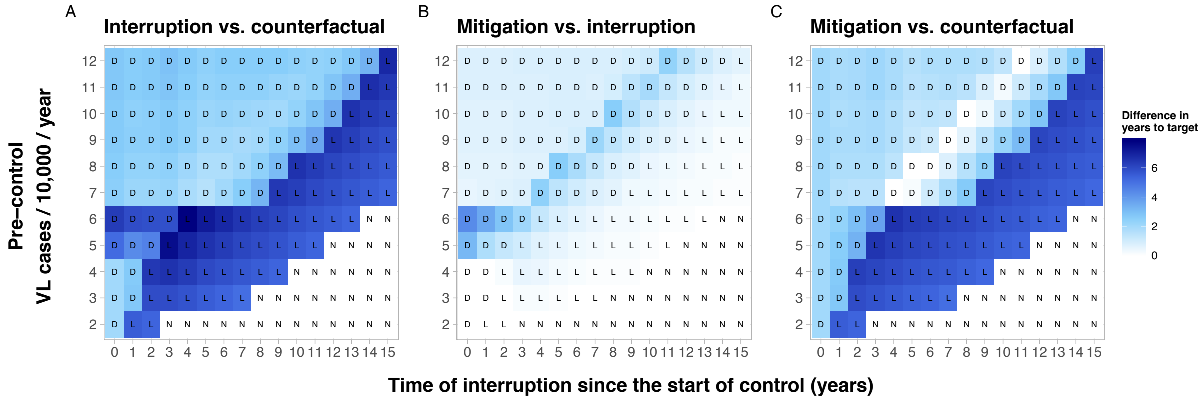

**Figure S11. Model E0: Heatmaps presenting the differences in time to achieving the VL elimination target for three combinations of scenarios (in years) with a 24-month interruption.** Panel A: interruption vs. counterfactual; Panel B: mitigation vs interruption, and Panel C: mitigation vs counterfactual. The letters D, L, and N correspond to the impact on the target when comparing the counterfactual scenario to the interruption scenario; delayed, lost, or not affected.

#### 5.4 Differences in cumulative VL incidence for 3 alternative durations of interruption

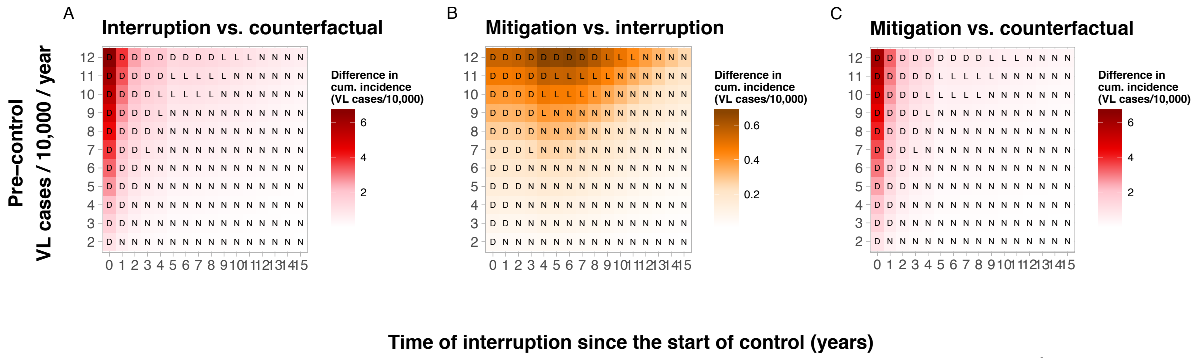

**Figure S12. Model E1: Heatmaps presenting the differences in cumulative incidence (VL cases/10,000) to achieving the VL elimination target for three combinations of scenarios with a 6-month interruption.** Panel A: interruption vs. counterfactual; Panel B: mitigation vs interruption, and Panel C: mitigation vs counterfactual. We use a different colour in panel B to indicate the finer scale relative to that depicted in panels A and C. The letters D, L, and N correspond to the impact on the target when comparing the counterfactual scenario to the interruption scenario; delayed, lost, or not affected.

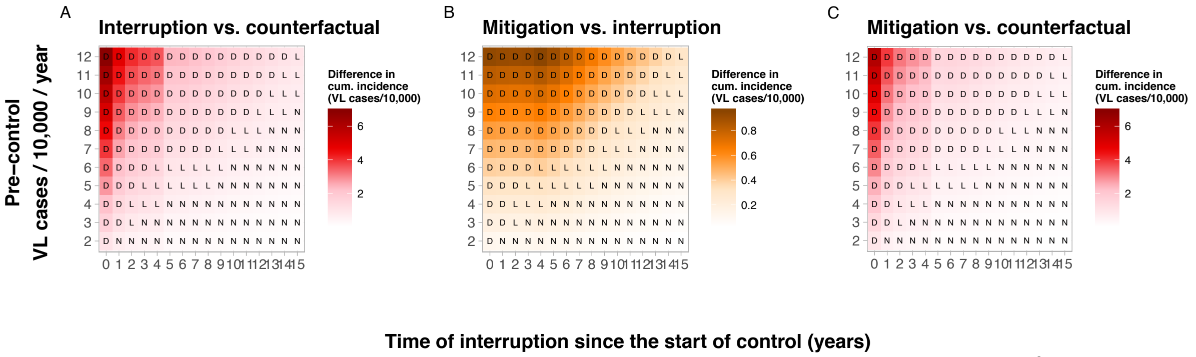

**Figure S13. Model E0: Heatmaps presenting the differences in cumulative incidence (VL cases/10,000) to achieving the VL elimination target for three combinations of scenarios with a 6-month interruption.** Panel A: interruption vs. counterfactual; Panel B: mitigation vs interruption, and Panel C: mitigation vs counterfactual. We use a different colour in panel B to indicate the finer scale relative to that depicted in panels A and C. The letters D, L, and N correspond to the impact on the target when comparing the counterfactual scenario to the interruption scenario; delayed, lost, or not affected.

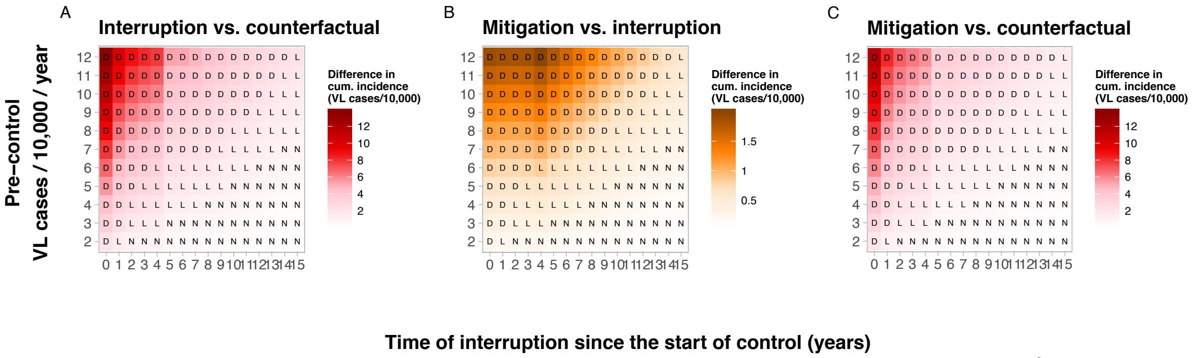

**Figure S14. Model E0: Heatmaps presenting the differences in cumulative incidence (VL cases/10,000) to achieving the VL elimination target for three combinations of scenarios with a 12-month interruption.** Panel A: interruption vs. counterfactual; Panel B: mitigation vs interruption, and Panel C: mitigation vs counterfactual. We use a different colour in panel B to indicate the finer scale relative to that depicted in panels A and C. The letters D, L, and N correspond to the impact on the target when comparing the counterfactual scenario to the interruption scenario; delayed, lost, or not affected.

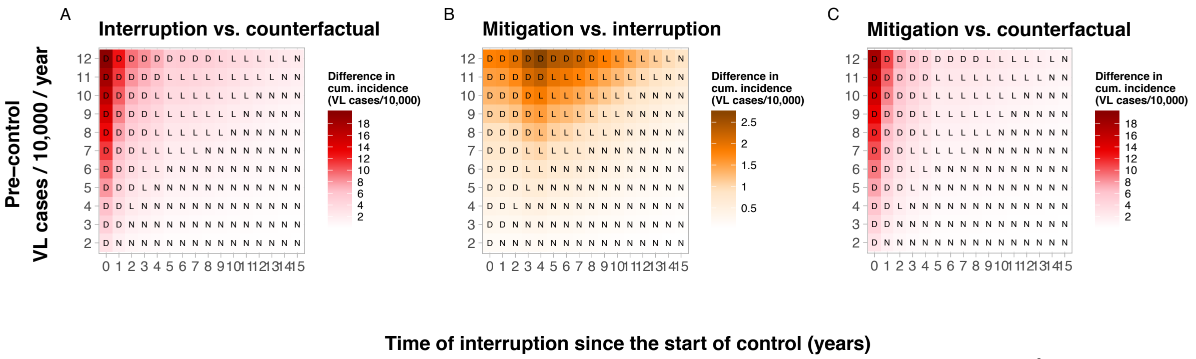

**Figure S15. Model E1: Heatmaps presenting the differences in cumulative incidence (VL cases/10,000) to achieving the VL elimination target for three combinations of scenarios with an 18-month interruption.**

Panel A: interruption vs. counterfactual; Panel B: mitigation vs interruption, and Panel C: mitigation vs counterfactual. We use a different colour in panel B to indicate the finer scale relative to that depicted in panels A and C. The letters D, L, and N correspond to the impact on the target when comparing the counterfactual scenario to the interruption scenario; delayed, lost, or not affected.

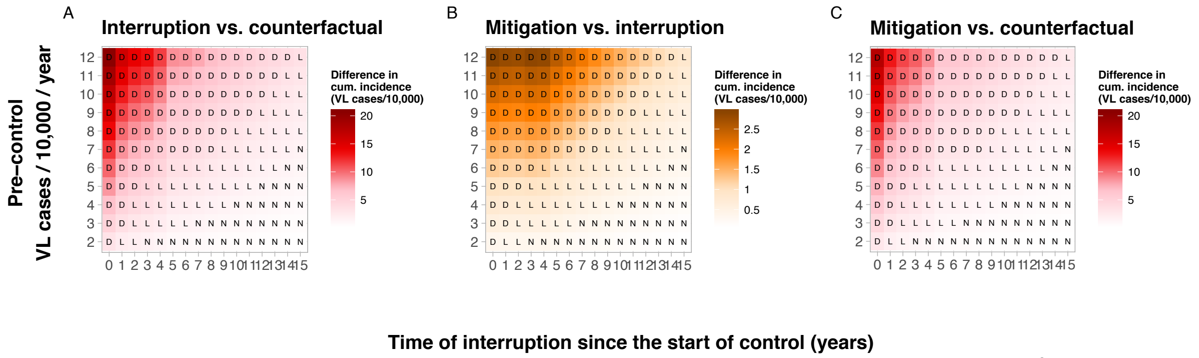

**Figure S16. Model E0: Heatmaps presenting the differences in cumulative incidence (VL cases/10,000) to achieving the VL elimination target for three combinations of scenarios with an 18-month interruption.** Panel A: interruption vs. counterfactual; Panel B: mitigation vs interruption, and Panel C: mitigation vs counterfactual. We use a different colour in panel B to indicate the finer scale relative to that depicted in panels A and C. The letters D, L, and N correspond to the impact on the target when comparing the counterfactual scenario to the interruption scenario; delayed, lost, or not affected.

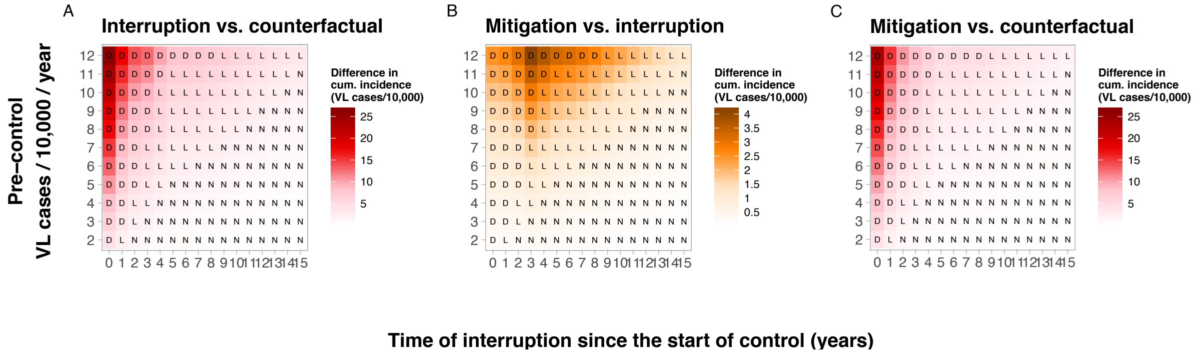

**Figure S17. Model E1: Heatmaps presenting the differences in cumulative incidence (VL cases/10,000) to achieving the VL elimination target for three combinations of scenarios with a 24-month interruption.** Panel A: interruption vs. counterfactual; Panel B: mitigation vs interruption, and Panel C: mitigation vs counterfactual. We use a different colour in panel B to indicate the finer scale relative to that depicted in panels A and C. The letters D, L, and N correspond to the impact on the target when comparing the counterfactual scenario to the interruption scenario; delayed, lost, or not affected.

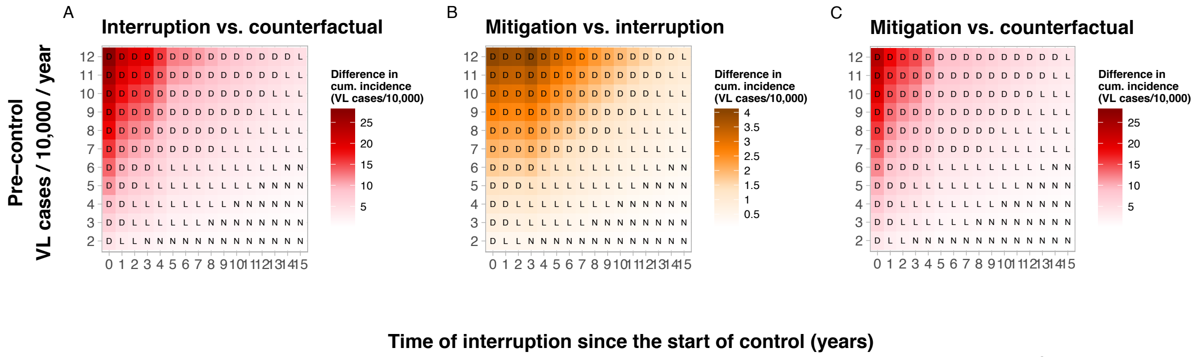

**Figure S18. Model E0: Heatmaps presenting the differences in cumulative incidence (VL cases/10,000) to**

**achieving the VL elimination target for three combinations of scenarios with a 24-month interruption.** Panel A: interruption vs. counterfactual; Panel B: mitigation vs interruption, and Panel C: mitigation vs counterfactual. We use a different colour in panel B to indicate the finer scale relative to that depicted in panels A and C. The letters D, L, and N correspond to the impact on the target when comparing the counterfactual scenario to the interruption scenario; delayed, lost, or not affected.

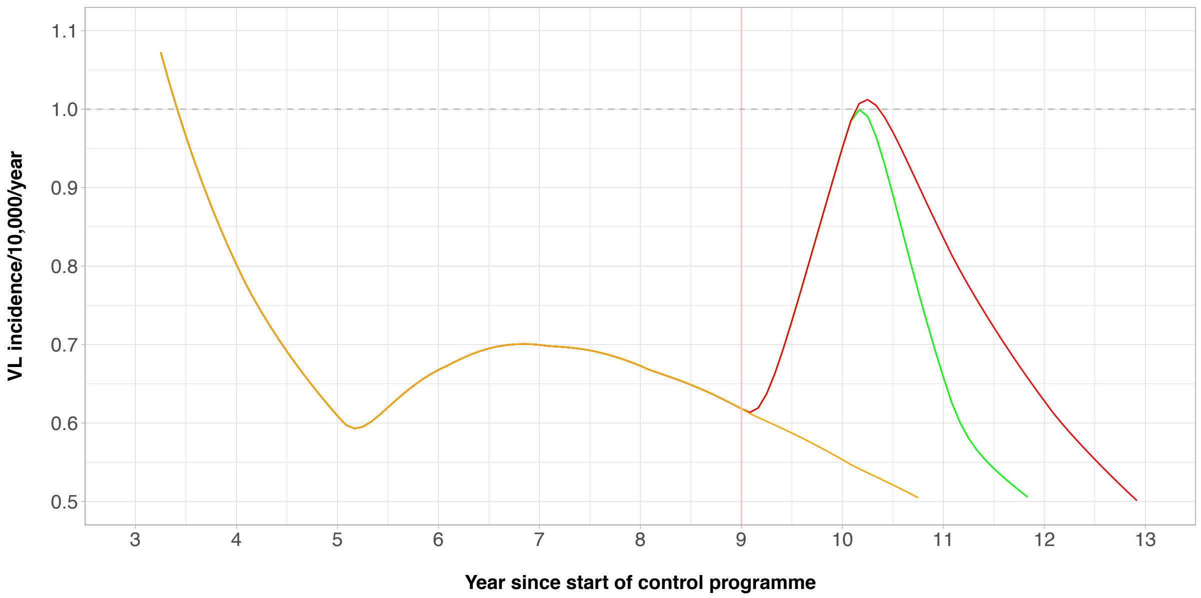

**Figure S19. Zoom in of VL incidence over time for a setting with a pre-control endemicity of 9/10,000/year with a 1-year interruption of the control programme 9 years after the start of the programme (Model E1).** The orange line represents the default scenario, the red line the interruption scenario, and the green line the mitigation scenario. The horizontal grey dashed line represents the 1/10,000/year elimination target. The pink vertical line represents the timing of the interruption.
